# Socioeconomic inequalities of Long COVID: findings from a population-based survey in the United Kingdom

**DOI:** 10.1101/2022.10.19.22281254

**Authors:** Sharmin Shabnam, Cameron Razieh, Hajira Dambha-Miller, Tom Yates, Clare Gillies, Yogini V Chudasama, Manish Pareek, Amitava Banerjee, Ichiro Kawachi, Ben Lacey, Eva JA Morris, Martin White, Francesco Zaccardi, Kamlesh Khunti, Nazrul Islam

## Abstract

**Objective:** To estimate the risk of Long COVID by socioeconomic deprivation and to further examine the socioeconomic inequalities in Long COVID by sex and occupational groups.

**Design:** We analysed data from the COVID-19 Infection Survey conducted by the Office for National Statistics between 26/04/2020 and 31/01/2022. This is the largest and nationally representative survey of COVID-19 in the UK and provides uniquely rich, contemporaneous, and longitudinal data on occupation, health status, COVID-19 exposure, and Long COVID symptoms.

**Setting:** Community-based longitudinal survey of COVID-19 in the UK.

**Participants:** We included 201,799 participants in our analysis who were aged between 16 and 64 years and had a confirmed SARS-CoV-2 infection.

**Main outcome measures:** We used multivariable logistic regression models to estimate the risk of Long COVID at least 4 weeks after acute SARS-CoV-2 infection by deciles of index of multiple deprivation (IMD) and adjusted for a range of demographic and spatiotemporal factors. We further examined the modifying effects of socioeconomic deprivation by sex and occupational groups.

**Results:** A total of 19,315 (9.6%) participants reported having Long COVID symptoms. Compared to the least deprived IMD decile, participants in the most deprived decile had a higher adjusted risk of Long COVID (11.4% vs 8.2%; adjusted OR: 1.45; 95% confidence interval [CI]: 1.33, 1.57). There were particularly significantly higher inequalities (most vs least deprived decile) of Long COVID in healthcare and patient facing roles (aOR: 1.76; 1.27, 2.44), and in the education sector (aOR: 1.62; 1.26, 2.08). The inequality of Long COVID was higher in females (aOR: 1.54; 1.38, 1.71) than males (OR: 1.32; 1.15, 1.51).

**Conclusions:** Participants living in the most socioeconomically deprived areas had a higher risk of Long COVID. The inequality gap was wider in females and certain public facing occupations (e.g., healthcare and education). These findings will help inform public health policies and interventions in adopting a social justice and health inequality lens.

## Introduction

The COVID-19 (Coronavirus Disease 2019) pandemic has led to an unprecedented public health crisis. Extensive research efforts have outlined the severe impact of COVID-related morbidity and mortality burden.^1–4^ A growing body of evidence also suggests that COVID-19 is a complex systemic disease that can leave a long-term impact to those affected.^5^ The National Institute for Health and Care Excellence (NICE) guidelines in the UK have included these ongoing symptoms under the umbrella term of “Long COVID”, comprising both “ongoing symptomatic COVID-19” (symptoms persisting for 4-12 weeks after the onset of acute infection) and “post COVID syndrome” (≥12 weeks after acute infection).^6^ These lingering symptoms are diverse in range and include both physical and psychological manifestations.^7,8^

According to the Office for National Statistics (ONS), an estimated 2 million people in the UK (3.1% of the population) are currently experiencing COVID-related symptoms persisting for more than four weeks, with 67% (1.2 million people) reporting that their daily activities had been adversely affected by prolonged symptoms, while 19% (376,000) of this group first had COVID-19 at least two years previously.^9^ The underlying mechanisms of Long COVID are still unclear;^8^ however studies report that these symptoms can occur even after having mild or asymptomatic SARS-CoV-2 infection. Given the extent of the long-term health risk, Long COVID has been identified as one of the priority areas for further research.^10^

Previous studies have found significantly higher risk of COVID-19 exposure, hospitalization, and mortality in the elderly, in ethnic minority populations, in people living in areas of lower socioeconomic status, and in those working in certain employment sectors (e.g., healthcare and frontline workers).^11–17^ The disproportionate impact of pandemic on people living in deprived areas may partly be due to having greater concentration of minority ethnic groups, higher prevalence of chronic medical conditions, occupational exposure, heavy reliance on public transport, crowded or multigenerational households, and limited access to healthcare.^18,19^ Occupation is particularly important because workplace setting can modify exposure risk (e.g., a higher exposure risk for public or client facing roles) as well as the effect of the exposure on various COVID-19 related outcomes.^20,21^

Such findings of differential exposure and consequently higher SARS-CoV-2 infection rates in certain vulnerable groups warrant the need to investigate whether such associations also exist in cases of Long COVID. Although sociodemographic and occupational inequality in Long COVID is currently largely unexplored, understanding this complex relationship is highly relevant in terms of assessing any unequal impact of the pandemic and adopting targeted and proportionate public health measures.^22^

Therefore, we undertook a study to estimate the risk of Long COVID by socioeconomic deprivation, independently of other potentially important predictors of Long COVID. We further examined the socioeconomic differentials in Long COVID by sex and occupational groups.

## Methods

### Data source

The study uses data from the COVID-19 infection survey (CIS). The CIS, conducted by the ONS and the University of Oxford, was approved by the South-Central Berkshire B Research Ethics Committee (20/SC/0195). It is a nation-wide longitudinal survey monitoring SARS-CoV-2 infection and immunity response in the UK.^23^ Private households were randomly selected from databases of addresses to ensure they reflect the UK population. Nose and throat swabs for Polymerase Chain Reaction (PCR) testing and blood samples for antibody testing were collected at regular intervals from individuals agreeing to participate. Participants also reported results of swab or blood tests that were collected and analysed elsewhere (for example, at general practices or home lateral flow tests). Participants also provided information about their demographic characteristics (age, sex, and ethnicity), occupation, and presence of long-term health conditions at each survey visit. Since February 3, 2021, the ONS included a section in the survey questionnaire pertaining to Long COVID symptoms and their severity.

The detailed protocol, survey design, and questionnaires of CIS are available online.^24–26^

### Study population

The data for this analysis were collected by the ONS CIS from April 26, 2020 to January 31, 2022. Participants were eligible for analysis if they were aged between 16 and 64 years at the first survey visit to reflect the working age population in UK.^27^

To assess the risk of Long COVID, we first restricted the analysis to survey participants with a confirmed SARS-CoV-2 infection and a corresponding date of first infection (see definitions below). Participants were excluded if they did not participate in the survey after February 3, 2021 (date when Long COVID questions were introduced in the survey) or if they did not participate at least 4 weeks of after their date of first infection.

### Definition of infection date and Long COVID

Participants were regarded as having Long COVID if they answered “yes” to: (1) having symptoms persisting for more than 4 weeks after their first SARS-CoV-2 infection which could not be explained by something else; or (2) having Long COVID symptoms that affected their day-to-day activities; or (3) having any specific Long COVID symptoms including pre-existing symptoms which were aggravated by a subsequent COVID-19 infection (**Supplementary Figure S1**).

COVID-19 cases were defined using both self-reported and ONS-conducted test results. In addition to the swab and blood test results (CIS or non-CIS) recorded at each visit, participants were also asked to report whether they suspected they had COVID-19 before the survey visit and the date of first suspected infection. We defined the first SARS-CoV-2 infection date (index date) as the earliest of the following: (1) date of first positive CIS PCR swab result; (2) date of first positive CIS blood sample result; (3) date of first positive non-CIS swab; (4) date of first positive non-CIS blood sample result; or (5) the self-reported date when the participant thought they had COVID-19.

Participants were followed from the first visit date occurring at least 4 weeks after the index date. The end of follow-up was defined as either: (1) the first visit date when the participant reported having Long COVID; or (2) the date of last follow-up visit by the end of the study period of the participant, if they did not report having Long COVID.

### Exposure

The primary exposure of interest was area-level socioeconomic deprivation measured using the area-based indices of Multiple Deprivation (IMD).^28^ The indices were developed using a combined relative measures of seven domains such as income, employment, health, education, crime, housing, and living environment. IMD is the official measure of deprivation in the UK. IMD scores and deciles were linked to the CIS data for each participant matched to their address and postcode. For our analysis, we have used IMD deciles where decile 1 represents the most deprived 10% of small areas and decile 10 represents the least deprived 10%.

### Covariates

Age was estimated from participants’ response during the first survey visit. Self-reported ethnicity was categorized as White or non-White due to small strata after further stratification (e.g., by IMD deciles, sex, regions). Ongoing long-term conditions (if the participants reported having any physical or mental health conditions excluding any long-lasting COVID-19 symptoms that lasted or expected to last for ≥1 year),^29^ household size,^30,31^ urban or rural residence, country of the respondents, and calendar time period of the index date (expressed as quarter of the year) were also included as covariates in the analysis. Household size of the participants was grouped into three categories: households of one person, two persons, and three persons or more.

We categorized participants’ occupations into four groups based on responses from each individual concerning whether their current job regularly involves in-person contact with patients or clients: (1) patient-facing healthcare workers; (2) non-patient facing healthcare workers; (3) patient or client-facing non-healthcare workers; (4) others (including workers in non-patient or client facing role, unemployed, unknown etc.). We adjusted for this variable in our main analysis.

We had complete data on all variables except self-reported on-going long-term conditions and occupation. Since participants’ data were collected at each survey visit, any missing records were imputed with the most recent valid data. We also excluded four occupational groups because of insufficient event counts by IMD deciles (fewer than 50) (**Supplementary Table S1**).

### Statistical Analysis

We generated descriptive statistics of demographic characteristics at the index date for the overall cohort, and for IMD decile 1 (most deprived) and decile 10 (least deprived).

We used multivariable logistic regression models to estimate the unadjusted and adjusted odds ratio (OR) and corresponding 95% confidence intervals (CI) of experiencing Long COVID symptoms by IMD deciles (using the least deprived group as the reference). We adjusted for age, sex, ethnicity, urban or rural location, comorbid conditions, household size, quarter of the year and country. We also included the logarithm of the follow-up time as an offset term. Confidence intervals were estimated using robust variance estimator.

Since the odds ratio only provides a relative risk for the group of interest compared to the reference group, we also estimated the absolute adjusted marginal risk from the regression models. We also conducted stratified analysis by sex and occupational groups to investigate whether there were any differences in risks within these categories.

### Sensitivity analysis

As a sensitivity analysis, we used multilevel logistic regression models, fitting random effects at country level to allow for the clustering of the data, adjusting for the same covariates. Since each of the four countries in the UK have used slightly different methods for measuring deprivation, we conducted another sensitivity analysis using only participants residing in England. We also carried out an additional sensitivity analysis using only the positive test results from PCR swab or blood sample (excluding self-reported data) to define the COVID-19 infection date.

All data management and statistical analyses were performed using Python version 3.6 and Stata MP Version 16.

## Results

During the study period, data were collected from a total of 535,634 participants, of whom 332,931 were between 16-64 years. Among them, 201,799 participants were eligible for our analysis (study flow chart available in **Supplementary Figure S2**).

Participants had a median follow-up duration of 214 days (interquartile rage [IQR]: 187-331 days) and median number of follow-up visits (at least 4 weeks after the index date) of 6 (IQR: 3-9).

**Table 1** reports the baseline (at the index date) demographic characteristics of the participants analysed, as well as a comparison between the most and the least deprived IMD deciles. Overall, the mean age of the participants was 45.1 (SD 12.9) years. Of them, 55.8% were female (n=112,683), 92.4% were White (n=186,547), and 79.6% were from urban areas (n=160,623). Most of the participants lived in a household of at least three persons (n=100,996, 50.0%), whereas 36.8% (n=74,264) and 13.2% (n=26,539) in two and one person households, respectively; 145,886 (72.3%) participants had a concurrent comorbidity and 12,934 (6.4%) were engaged in a healthcare occupation with a patient facing role.

**Table 1:** Baseline characteristics of the cohort.

| Characteristics | IMD<br>1 (most deprived)<br>(N=9,483) | IMD<br>10 (least deprived)<br>(N= 27,113) | Overall<br>(N=201,799) |
| --- | --- | --- | --- |
| Age, mean (SD)) | 43.8 (13.0) | 46.3 (12.7) | 45.1 (12.9) |
| Age, (median (IQR)) | 45.0 (33.0-55.0) | 48.0 (39.0-57.0) | 47.0 (36.0-56.0) |
| Sex, n (%) |  |  |  |
| Male | 4,048 (42.7) | 12,234 (45.1) | 89,116 (44.2) |
| Female | 5,435 (57.3) | 14,879 (54.9) | 112,683 (55.8) |
| Ethnicity, n (%) |  |  |  |
| White | 8,600 (90.7) | 25,679 (94.7) | 186,547 (92.4) |
| Non-White | 883 (9.3) | 1,434 (5.3) | 15,252 (7.6) |
| Rural/urban, n (%) |  |  |  |
| Urban | 9,211 (97.1) | 22,462 (82.8) | 160,623 (79.6) |
| Rural | 272 (2.9) | 4,651 (17.2) | 41,176 (20.4) |
| Household size in persons, n (%) |  |  |  |
| 1 | 2,081 (21.9) | 2,299 (8.5) | 26,539 (13.2) |
| 2 | 3,252 (34.3) | 89,68 (33.1) | 74,264 (36.8) |
| >=3 | 4,150 (43.8) | 15,846 (58.4) | 100,996 (50.0) |
| Any comorbid conditions, n (%) |  |  |  |
| No | 5,484 (57.8) | 20,855 (76.9) | 145,886 (72.3) |
| Yes | 3,999 (42.2) | 6,258 (23.1) | 55,913 (27.7) |
| Country, n (%) |  |  |  |
| England | 7,989 (84.2) | 22,668 (83.6) | 171,602 (85.0) |
| Scotland | 777 (8.2) | 2,272 (8.4) | 15,253 (7.6) |
| Wales | 498 (5.3) | 1,233 (4.5) | 9,232 (4.6) |
| Northern Ireland | 219 (2.3) | 940 (3.5) | 5,712 (2.8) |
| Occupational role |  |  |  |
| Non-healthcare patient/client facing | 1,943 (20.5) | 5,324 (19.6) | 42,013 (20.8) |
| Healthcare non-patient/client facing | 326 (3.4) | 1,081 (4.0) | 7,417 (3.7) |
| Healthcare patient/client facing | 624 (6.6) | 1,675 (6.2) | 12,934 (6.4) |
| Other or Unknown | 6,590 (69.5) | 19,033 (70.2) | 139,435 (69.1) |

IMD: Index of Multiple Deprivation; SD: standard deviation; IQR: interquartile range.

Compared to the least deprived, participants in the most deprived decile had a lower mean age (43.8 vs 46.3 years), lower proportion of males (42.7% vs 45.1%) and lower proportion from White ethnic group (90.7% vs 94.7%), and had a higher proportion of participants from urban areas (97.1% vs 82.8%) and one person households (21.9% vs 8.5%). Participants residing in the least deprived areas had a substantially lower prevalence of any on-going health conditions (23.1%) compared to those in the most deprived (42.2%). The distribution of country of residence and occupational roles were comparable between the most and the least deprived areas (**Table 1**).

One in ten (9.6%; n=19,315) participants reported having Long COVID symptoms that persisted 4 or more weeks after the acute infection. The prevalence of Long COVID varied by sex and IMD decile: it was higher in females (n= 11,875, 11.8%) than males (n=7,440, 9.1%) and in participants residing in the most deprived tenth of areas (n=1,229, 13.0%) versus those who resided in the least deprived tenth of areas (n=2,188, 8.1%) (**Supplementary Table S2**). The prevalence of Long COVID by IMD and occupational groups showed similar trends (**Supplementary Table S3**).

### Adjusted odds ratio of Long COVID by IMD deciles

The adjusted odds of having Long COVID was greater in more deprived neighbourhoods (**Supplementary Figure S3**). After adjusting for potential covariates, the odds of Long COVID were 46% higher (OR: 1.46; 1.34, 1.59) in the most deprived decile (compared to the least). The odds of long COVID were similar to those of the main analysis when the participants residing in England were analysed separately (OR: 1.41; 1.29, 1.54) in a sensitivity analysis. The trend was also consistent (OR: 1.46; 1.31, 1.63) in a sensitivity analysis using random effects at country level. Results from an additional sensitivity analysis excluding the participants who only self-reported having COVID-19 also showed comparable results (OR: 1.52; 1.37, 1.69) (**Supplementary Tables S4-S6**).

When stratified by sex, the results were generally consistent in terms of deprivation-specific trends, with a higher level of inequality among females (OR: 1.56; 1.40, 1.73) than males (OR: 1.32; 1.15, 1.51) in the most deprived decile compared to the least deprived decile (**Figure 1**).

**Figure 1.**
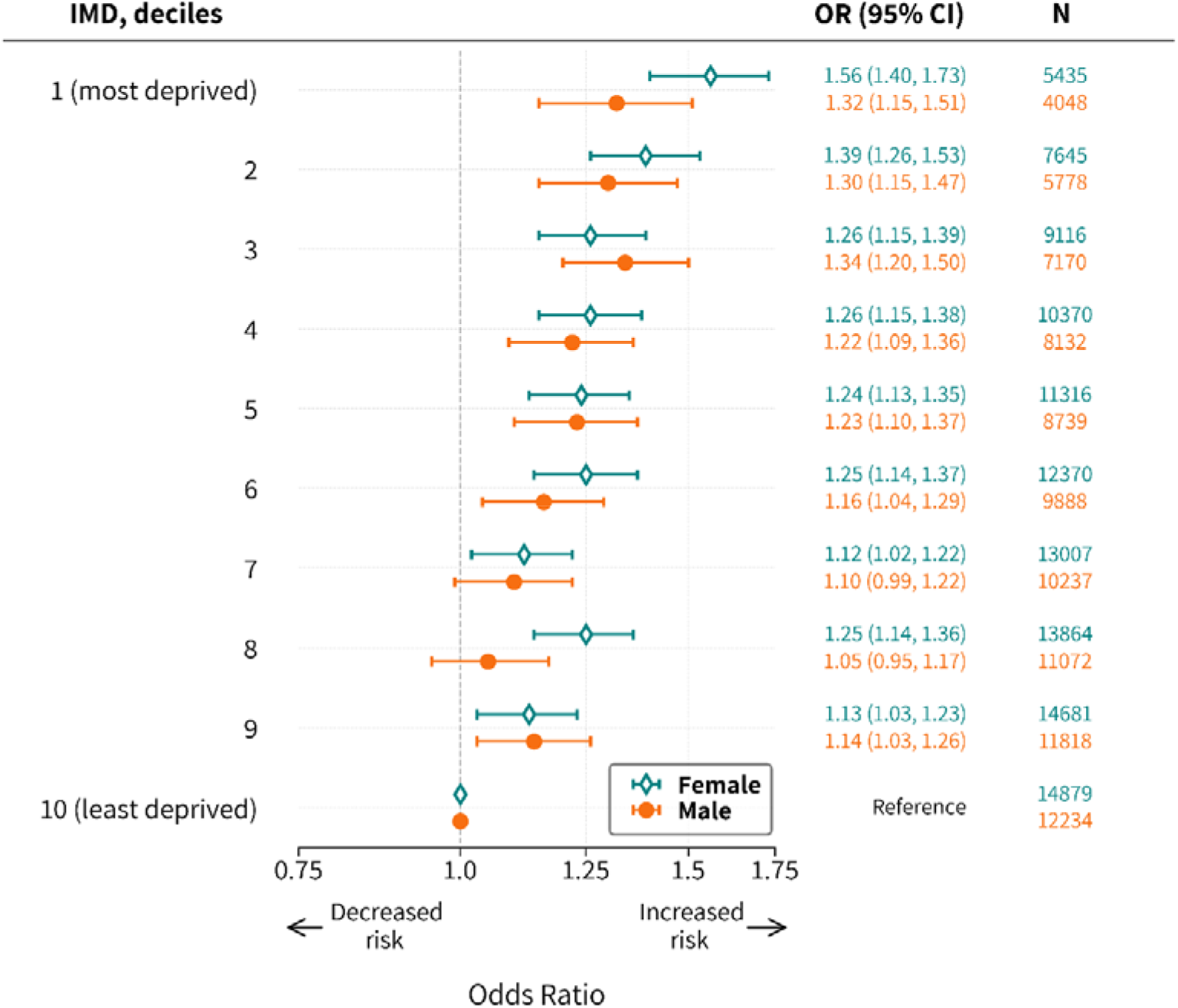
Association between deprivation and experiencing Long COVID at least 4 weeks after having COVID-19, stratified by sex. Estimates adjusted for age, ethnicity, urban/rural, comorbid conditions, household size, country, quarter of the year, healthcare and patient/client-facing nature of the job in the multivariable logistic regression model using the logarithm of the follow-up time as an offset term.

We also investigated whether the risk of Long COVID was modified by the occupation of participants and found large variations in different occupational groups. For example, compared to the least deprived decile, the adjusted odds ratio of having Long COVID was significantly higher in participants from the most deprived decile for those working in health care and patient facing role (OR: 1.76; 1.27, 2.44), teaching and education roles (OR: 1.68; 1.31, 2.16), overall health care role (OR: 1.65; 1.27, 2.14), hospitality sector (OR: 1.58; 1.01, 2.46), and civil service (OR: 1.44; 1.01, 2.05) (**Figure 2**). Our analysis showed no statistically significant association in the relative risk of Long COVID for those working in other employment sectors such as social care, retail, and manufacturing or construction sector.

**Figure 2.**
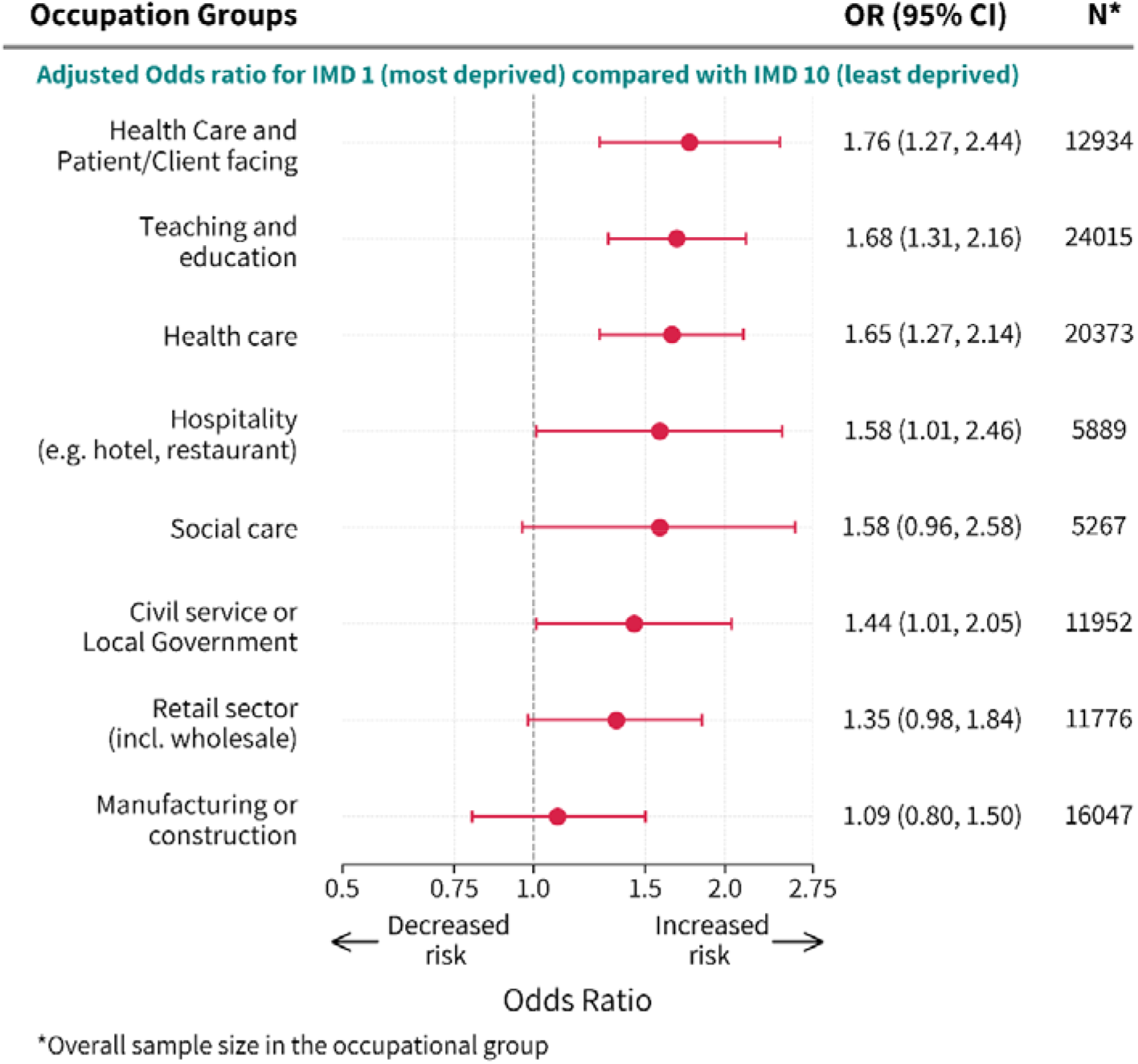
Association between deprivation and experiencing Long COVID at least 4 weeks after having COVID-19, stratified by occupational groups. Estimates adjusted for age, sex, ethnicity, urban/rural, comorbid conditions, household size, country and quarter of the year in the multivariable logistic regression model using the logarithm of the follow-up time as an offset term.

### Adjusted absolute risks of Long COVID by IMD deciles

Overall, the adjusted prevalence of people reporting any Long COVID symptoms at least 4 weeks after having COVID-19 was higher in the most deprived decile (11.4%; 10.8, 12.1) than the least deprived decile (8.2%; 7.9, 8.6) (**Table 2**). The adjusted prevalence also increased with increasing levels of deprivation.

**Table 2:** Proportion of participants experiencing Long COVID at least 4 weeks after having COVID-19, by IMD deciles.

| IMD, deciles | Unadjusted<br>% (95% CI) | Adjusted*<br>% (95% CI) | Sex Stratified** |  |
| --- | --- | --- | --- | --- |
|  |  |  | Male<br>% (95% CI) | Female<br>% (95% CI) |
| **1 (most deprived)** | 12.71 (12.00, 13.42) | 11.40 (10.75, 12.06) | 9.35 (8.44, 10.26) | 13.00 (12.07, 13.92) |
| **2** | 11.28 (10.72, 11.84) | 10.68 (10.14, 11.22) | 9.24 (8.47, 10.01) | 11.81 (11.06, 12.56) |
| **3** | 10.49 (9.99, 10.98) | 10.27 (9.79, 10.76) | 9.47 (8.76, 10.18) | 10.91 (10.24, 11.59) |
| **4** | 10.00 (9.54, 10.45) | 9.93 (9.48, 10.38) | 8.72 (8.08, 9.36) | 10.89 (10.26, 11.51) |
| **5** | 9.89 (9.46, 10.33) | 9.87 (9.44, 10.30) | 8.80 (8.18, 9.42) | 10.72 (10.12, 11.32) |
| **6** | 9.55 (9.14, 9.95) | 9.74 (9.32, 10.15) | 8.36 (7.79, 8.94) | 10.82 (10.24, 11.41) |
| **7** | 8.83 (8.45, 9.21) | 9.01 (8.62, 9.40) | 7.97 (7.42, 8.52) | 9.84 (9.29, 10.38) |
| **8** | 9.21 (8.83, 9.58) | 9.41 (9.02, 9.79) | 7.66 (7.14, 8.17) | 10.80 (10.25, 11.35) |
| **9** | 8.99 (8.63, 9.35) | 9.18 (8.81, 9.54) | 8.23 (7.71, 8.75) | 9.92 (9.41, 10.43) |
| **10 (least deprived)** | 8.07 (7.74, 8.41) | 8.23 (7.89, 8.57) | 7.33 (6.84, 7.81) | 8.94 (8.46, 9.42) |

\*Adjusted for age, sex, ethnicity, urban/rural, comorbid conditions, household size, healthcare and patient/client facing nature of the job, and country in the logistic regression model using logarithm of the follow-up time as an offset term.
\*\*Adjusted for age, ethnicity, urban/rural, comorbid conditions, household size, healthcare and patient/client-facing nature of the job, and country in the logistic regression model using logarithm of the follow-up time as an offset term.

In males, the adjusted prevalence of participants reporting any Long COVID symptoms ranged from 7.3% (6.8, 7.8) in the least deprived to 9.4% (8.4, 10.3) in the most deprived decile. In females, the range was from 8.9% (8.5, 9.4) to 13.0% (12.1, 13.9) in the least and the most deprived deciles, respectively. The risk in females in the least deprived decile was comparable to that in males living the most deprived decile (**Table 2**).

When stratified by occupational groups, the absolute risk was always higher in participants residing in the most deprived areas than those in the least deprived areas. However, the adjusted prevalence varied substantially even when the deprivation level was the same. For example, in the most deprived decile, the prevalence ranged from 10.0% (7.7, 12.3) in the manufacturing or construction sector to 14.6% (12.1, 17.2) in teaching and education sector. In contrast, the variability within the least deprived decile was relatively smaller, and ranged from 8.3% (6.1, 10.5) in the hospitality sector to 9.5% (8.5, 10.5) in teaching and education sector (**Table 3**).

**Table 3:** Adjusted proportions of participants experiencing Long COVID at least 4 weeks after having COVID-19, by occupational groups and IMD deciles.

| Occupational groups | IMD 1 (most deprived);<br>adjusted proportions<br>(95% CI) | IMD 10 (least deprived);<br>adjusted proportions<br>(95% CI) |
| --- | --- | --- |
| Teaching and education | 14.64 (12.11, 17.18) | 9.48 (8.47, 10.48) |
| Health care and patient/client facing | 13.94 (11.04, 16.83) | 8.75 (7.32, 10.18) |
| Health care | 13.14 (10.89, 15.39) | 8.66 (7.56, 9.76) |
| Civil service or local government | 12.32 (9.32, 15.32) | 9.05 (7.56, 10.53) |
| Social care | 12.31 (8.80, 15.83) | 8.36 (5.84, 10.87) |
| Hospitality | 12.24 (8.91, 15.58) | 8.28 (6.06, 10.50) |
| Retail sector | 12.04 (9.71, 14.38) | 9.36 (7.67, 11.06) |
| Manufacturing or construction | 10.02 (7.70, 12.34) | 9.27 (7.95, 10.58) |

Adjusted for age, sex, ethnicity, urban/rural, comorbid conditions, household size, and country in the logistic regression model using logarithm of the follow-up time as an offset term.

## Discussion

In this study, we investigated the risk of Long COVID in a large nationally representative community-based survey of over 200,000 working-age adults in successive waves of the COVID-19 pandemic and report that the risk of experiencing such symptoms is strongly associated with the area level of deprivation of the participants. Results from our analysis show that the odds of experiencing Long COVID are 45% higher on average for participants from the most deprived areas (IMD decile 1) compared to those in the least deprived areas (IMD decile 10). Our findings are robust after controlling for baseline demographic factors, household size, time of the year, comorbidity, and follow-up duration. The probability of persistent symptoms after at least 4 weeks was also the lowest in the least deprived population and increased in an almost dose-response fashion with increasing levels of deprivation, both in males and females. Females also exhibited an elevated risk of developing Long COVID after SARS-CoV-2 infection, compared to males across all the IMD deciles.

We also found that these associations varied widely by the occupational groups of the participants. Stratified analysis showed that those living in the most deprived areas and working in healthcare (both patient facing and non-patient facing roles) and in the teaching/education sectors had the highest likelihoods (76%, 64%, and 62%, respectively) of reporting Long COVID symptoms compared to the least deprived group, while no significant association was observed in other occupational groups. This indicates that socioeconomic disparities in risk of Long COVID are wider in certain occupational groups, and that these disparities cannot be explained by differential exposure to SARS-CoV-2 infection.

### Findings in context

To our knowledge, this is the first study to quantify the association between Long COVID symptoms by socioeconomic status and occupational groups. Previous research reported that people from lower socioeconomic backgrounds had worse COVID-19 outcomes in terms of infection severity, hospitalisation, and mortality. Our results suggest that the unequal effects of the COVID-19 pandemic on the most socioeconomically vulnerable population extends beyond recovery from acute illness. Only a few studies have examined the risk factors of Long COVID, and found that female sex, age, smoking, body mass index, and comorbidities are strong predictors.^32–38^ A cohort study of 4,182 COVID-19 patients showed that those who had more than five symptoms during the initial phase of acute illness were 3.5 times more likely to develop Long COVID.^33^ Another observational study reported a significant association between severe COVID-19 and persistent symptoms suggesting that the disease severity might lead to higher likelihood of Long COVID.^39^ One study^33^ found no significant variation within different socioeconomic groups while another^34^ reported that deprivation was associated with having Long COVID-19. However, none of the prior studies explored the intersectional inequalities (sex, deprivation, and occupation) of Long COVID risk as we did in our study.

Occupational groups such as healthcare, transport, retail, and social care workers have also been unequally affected by the COVID-19 pandemic.^13,14^ The heightened risk could be due to more frequent exposure to infections, lack of personal protective equipment, lack of provision to work from home, higher usage of public transport, not being able to take time off from work, lack of proper ventilation, mask wearing and physical distancing at work.^40^ The occupational inequalities resulted in higher SARS-CoV-2 exposure, infection, and mortality of frontline and essential workers compared to general population. However, studies regarding Long COVID and occupation are sparse. A recent study reported that participants working in healthcare had a higher probability of having long-term sequelae of COVID-19.^34^ The adverse effect of the pandemic on the occupational health of doctors have led the British Medical Association to insist on recognising Long COVID as an occupationally acquired disease.^41^ However, to our knowledge, no previous studies have investigated Long COVID among a wide range of occupational groups.

We have demonstrated that the risk of prolonged symptoms and illness related to COVID-19 is not homogenous across the intersections of work sectors and deprivation levels. We have found that the socioeconomic inequality, i.e., the difference between the most and the least deprived populations, was particularly higher among females and those employed in healthcare or teaching/education sector. Our findings are not directly comparable in the context of COVID-19 because no previous study has conducted such detailed analysis. However, our results are consistent with pre-pandemic research on other health conditions suggesting that workers with lower socioeconomic status have poorer health outcomes and higher premature mortality than those with higher socioeconomic position but a similar occupation.^42–46^ Poor working and housing conditions (including living in dense, poorly ventilated multigenerational households) and unhealthier lifestyle behaviour (poor nutrition, heavy alcohol consumption, smoking etc.) are among the well-established factors associated with the overall burden of ill health among socioeconomically deprived workers.^43^ Previous evidence, together with the results from this study, suggest that inequalities in Long COVID cannot be viewed in isolation without considering the role of occupation in a gender-blind manner.

### Implications for policy and practice

This study provides insights into the heterogeneous degree of inequality by deprivation, sex and occupation with regards to Long COVID. This indicates the need for a diverse range of public health interventions after recovery from COVID (treatment and/or rehabilitation) across multiple intersecting social dimensions as well as data for reducing disproportionate impact on these populations in any future waves of the pandemic. Hence, our findings highlight the necessity of managing post-recovery of COVID-19 with a health equity lens. These include assessing the differentials in Long COVID diagnosis, health-seeking behaviour, and follow-up after recovery across the intersections of sex, occupation, and socioeconomic circumstances of people. The assessment will help inform health policy in identifying the most vulnerable sub-groups of populations so that more focused efforts are given, and proportional allocation of resources are implemented to facilitate the reduction of health inequalities.

Most studies on health inequality have predominantly taken unitary approaches (focusing on sex, ethnicity or socioeconomic status separately) and rarely explored the impact of intersectional inequality on population health.^47–49^ However, the inequalities shown in this study shows that such approach can provide more precise identification of risks and be relevant to other diseases and beyond the pandemic.

### Strengths and limitations

The study has several key strengths. We have used the COVID-19 Infection Survey for our analysis which is the largest community-based longitudinal survey of COVID-19 in the UK. The survey provides rich, contemporaneous, and longitudinal data on socio-economic factors, occupation, health status, COVID-19 exposure, and Long COVID symptoms. The survey design ensures participation from representative households across the UK through random sampling to obtain direct population-level estimates of Long COVID.

Another strength of this study is that we have examined Long COVID in both asymptomatic and symptomatic COVID-19 patients. The participants in this study had their nose and throat swab collected at each visit. Consequently, the survey captured Long COVID data among the subgroup of the population who would not have been tested due to lack of COVID-19 symptoms. Our study is also strengthened by the usage of a consistent definition of Long COVID thought out the survey period.

We have also adjusted for a range of covariates in the model to estimate the independent effects of the IMD on the respective outcomes. We further examined intersectional inequality by estimating the differential risks of Long COVID within sex and occupational groups. However, our study has some limitations. First, IMD is an ecological measure; therefore, these findings may not be interpreted at individual participant level. Second, Long COVID symptoms are diverse in nature^7^ and our study may not have captured the full range of symptoms experienced by the participants. However, the 21 specific symptoms of Long COVID included in the survey questionnaire are selected based on a comprehensive review of the existing body of evidence and encompasses the most common types of neurological, psychological, and physiological symptoms reported by patients.^37,50^ Furthermore, the survey questionnaire allows participants to self-classify themselves as having Long Covid without the need to be experiencing the 21 specific symptoms listed on the questionnaire.

Third, Long COVID symptoms and existence of any chronic conditions were self-reported. Therefore, we could not rule out the possibility that some of the reported symptoms may not be directly caused by COVID-19. Furthermore, to address the possibility of a differential attribution of long COVID symptoms depending on the time from acute infection to the survey visit date, we controlled for follow-up time in our analyses.

Fourth, as this is an observational analysis, a causal relationship between socioeconomic deprivation and the risks of Long COVID cannot be established. Our results focus on the unequal long-term burden of COVID across the deprived communities in the UK and therefore the results we found only measure the association of the two in the aggregated area level.

Our research also could not examine the time to remission of Long COVID symptoms due to lack of granular longer term follow-up data. Future studies should fill this gap in the literature, especially if the time to remission varies by occupation and deprivation. Future research could also explore additional set of symptoms that are yet to be reported and recognised as Long COVID symptoms.

## Conclusion

Our study demonstrates that people from the most socioeconomically deprived populations have the highest risk of long COVID symptoms, and this inequality is independent of differences in the risk of initial infection. The trend was consistent within different sex and occupational groups, but the disparity gap was heterogeneous, with a particularly higher level of inequality in participants who are female, working in health care, and in teaching and education sector. Future health policy recommendations should incorporate the multiple dimensions of inequality, such as sex, deprivation, and occupational groups when considering the treatment and management of Long COVID.

## Data Availability

The data from the Office of National Statistics COVID-19 Infection Survey (CIS) can be accessed only by ONS accredited researchers (AR) through the Secure Research Service (SRS). Researchers can apply for accreditation through the Research Accreditation Service and will need approval to access CIS data. For further details see: https://www.ons.gov.uk/aboutus/whatwedo/statistics/requestingstatistics/secureresearchservice.

## Declarations

### Disclaimer

The opinions expressed within this report are solely the authors’ and do not necessarily reflect the opinions of the organisations/entities the authors are employed by and/or affiliated with.

“This work was produced using statistical data from the Office for National Statistics (ONS). The use of the ONS statistical data in this work does not imply the endorsement of the ONS in relation to the interpretation or analysis of the statistical data. This work uses research datasets which may not exactly reproduce National Statistics aggregates.”

### Ethics Approval

The ONS COVID-19 Infection Survey (CIS) was approved by the South-Central Berkshire B Research Ethics Committee (Ethics Ref: 20/SC/0195). The study was assessed using the National Statistician’s Data Ethics Advisory Committee (NSDEC) ethics self-assessment tool, and the committee confirmed that no further ethical consideration was required.

### Data availability statement

The data from the Office of National Statistics COVID-19 Infection Survey (CIS) can be accessed only by ONS accredited researchers (AR) through the Secure Research Service (SRS). Researchers can apply for accreditation through the Research Accreditation Service and will need approval to access CIS data. For further details see: https://www.ons.gov.uk/aboutus/whatwedo/statistics/requestingstatistics/secureresea rchservice.

### Guarantor

NI.

### Contributorship

NI and KK conceived and designed the study, obtained the funding, developed the statistical methodology, and managed and coordinated research activity. SS carried out the data preparation, analyses and data visualization. SS drafted the first version of the manuscript. NI contributed significant edits and input for the draft manuscript. All authors contributed to reviewing the manuscript and interpreting the findings. All authors have approved the final published version.

## Acknowledgements

We are grateful to Dr. Vahé Nafilyan and Daniel Ayoubkhani from the Office for National Statistics for their contributions to the analysis and very helpful feedback on earlier versions of the manuscript.

