## Supplementary Material for "Socioeconomic inequalities of Long COVID: findings from a population-based survey in the United Kingdom"

Figures 2

Figure S1: Long COVID symptoms in the survey questionnaire. 2

Figure S2: Flow chart for the analysis of Long COVID symptoms. 3

Figure S3: Association between deprivation and experiencing Long COVID at least 4 weeks after having COVID-19. 4

Tables 5

Table S1: Occupation groups listed in the survey questionnaire. 5

Table S2: Number of participants experiencing Long COVID at least 4 weeks after having COVID-19, by IMD deciles. 6

Table S3: Number of participants experiencing Long COVID at least 4 weeks after having COVID-19 by IMD deciles and occupational groups. 7

Table S4: Association between deprivation and experiencing Long COVID at least 4 weeks after having COVID-19, for participants residing in England. 8

Table S5: Association between deprivation and experiencing Long COVID at least 4 weeks after having COVID-19, using random-effects at country level 9

Table S6: Association between deprivation and experiencing Long COVID at least 4 weeks after having COVID-19, excluding those who only had self-reported COVID-19 results. 10

### Figures

#### Figure S1: Long COVID symptoms in the survey questionnaire.

5. Would you describe yourself as having "long COVID", that is, you are still experiencing symptoms more than 4 weeks after you first had COVID-19, that are not explained by something else? ☐ Yes ☐ No

*If yes*: (a) Does this reduce your ability to carry-out day-to-day activities compared with the time before you had COVID-19? (*select one*) ☐ Yes, a lot ☐Yes, a little ☐ Not at all

(b) Have you had any of the following symptoms as part of your experience of long COVID? Please include any pre-existing symptoms which long COVID has made worse (answer Yes or No for each one)

| | | | | | | | | |
|---|---|---|---|---|---|---|---|---|
| Fever (including high temperature) | ☐ Yes | ☐ No | Headache | ☐ Yes | ☐ No | Muscle ache | ☐ Yes | ☐ No |
| Weakness/tiredness | ☐ Yes | ☐ No | Nausea/vomiting | ☐ Yes | ☐ No | Abdominal pain | ☐ Yes | ☐ No |
| Diarrhoea | ☐ Yes | ☐ No | Loss of appetite or eating less than usual | ☐ Yes | ☐ No | Loss of taste | ☐ Yes | ☐ No |
| Loss of smell | ☐ Yes | ☐ No | Sore throat | ☐ Yes | ☐ No | Cough | ☐ Yes | ☐ No |
| Shortness of breath | ☐ Yes | ☐ No | Chest pain | ☐ Yes | ☐ No | Palpitations | ☐ Yes | ☐ No |
| Vertigo/dizziness | ☐ Yes | ☐ No | Worry/anxiety | ☐ Yes | ☐ No | Low mood/not enjoying anything | ☐ Yes | ☐ No |
| More trouble sleeping than usual | ☐ Yes | ☐ No | Memory loss or confusion | ☐ Yes | ☐ No | Difficulty concentrating | ☐ Yes | ☐ No |
| Runny nose/ sneezing | ☐ Yes | ☐ No | Noisy breathing (wheezing) | ☐ Yes | ☐ No | | | |

Reference: https://www.ndm.ox.ac.uk/covid-19/covid-19-infection-survey/case-record-forms?d3742abe-11b5-11ed-ab39-06d04f560572.

**Figure S2: Flow chart for the analysis of Long COVID symptoms.**

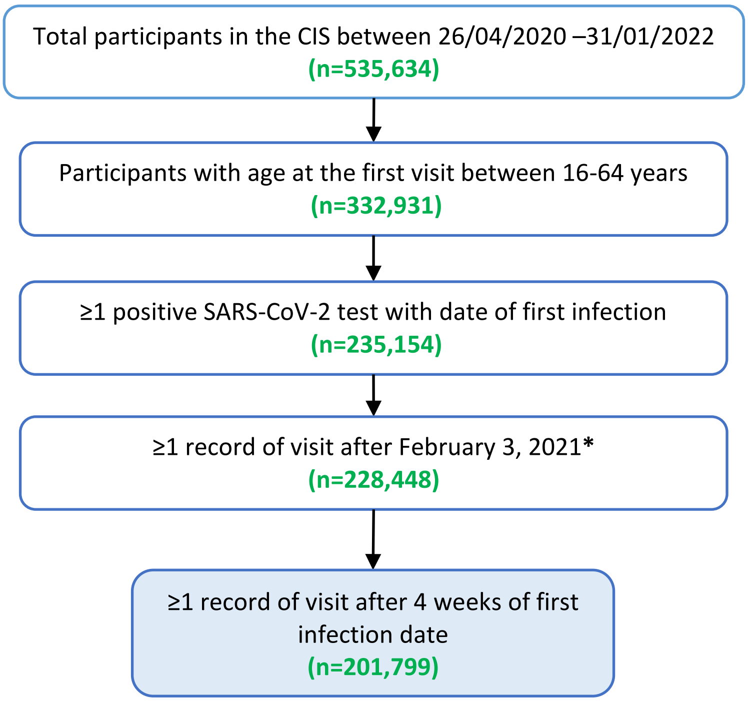


*February 3, 2021 was the date the Long COVID questionnaire was introduced in the ONS CIS survey.

**Figure S3: Association between deprivation and experiencing Long COVID at least 4 weeks after having COVID-19.**

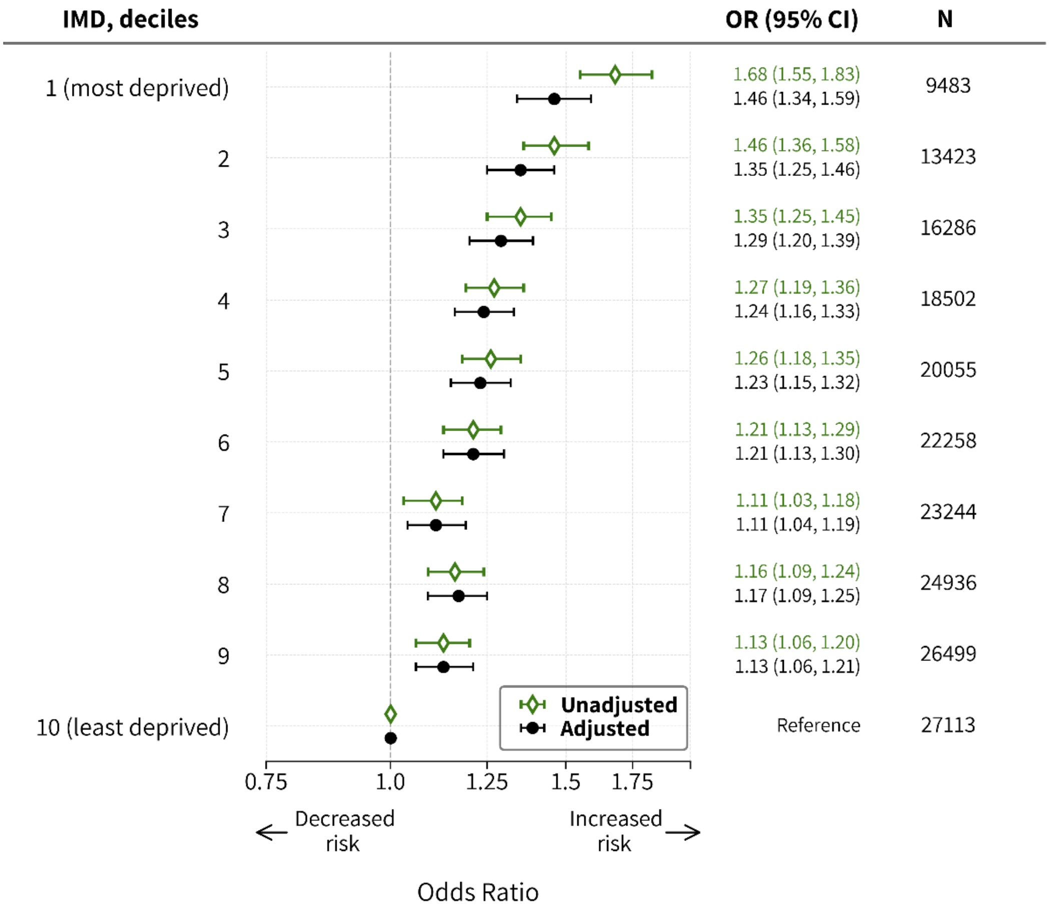


Estimates adjusted for age, sex, ethnicity, urban/rural, comorbid conditions, household size, healthcare and country in the logistic regression model using logarithm of the follow-up time as an offset term.

### Tables

**Table S1: Occupation groups listed in the survey questionnaire.**

| Occupation groups | Included in the analysis |
|---|---|
| Teaching and education | Yes |
| Health care | Yes |
| Manufacturing or construction | Yes |
| Financial services incl. insurance | No |
| Civil service or Local Government | Yes |
| Information Technology and communication | No |
| Retail sector (incl. wholesale) | Yes |
| Transport (incl. storage, logistic) | No |
| Hospitality (e.g., hotel, restaurant) | Yes |
| Social care | Yes |
| Arts, Entertainment or Recreation | No |
| Food production, agriculture, farming | No |
| Personal services (e.g., hairdressers) | No |
| Armed forces | No |
| Other employment sector | No |

**Table S2: Number of participants experiencing Long COVID at least 4 weeks after having COVID-19, by IMD deciles.**

| IMD, deciles | Number of participants | Number of participants experiencing Long COVID, n (%) |
|---|---|---|
| **1 (most deprived)** | 9483 | 1229 (12.96) |
| **2** | 13423 | 1534 (11.43) |
| **3** | 16286 | 1705 (10.47) |
| **4** | 18502 | 1851 (10.0) |
| **5** | 20055 | 1986 (9.9) |
| **6** | 22258 | 2109 (9.48) |
| **7** | 23244 | 2048 (8.81) |
| **8** | 24936 | 2291 (9.19) |
| **9** | 26499 | 2374 (8.96) |
| **10 (least deprived)** | 27113 | 2188 (8.07) |

**Table S3: Number of participants experiencing Long COVID at least 4 weeks after having COVID-19 by IMD deciles and occupational groups.**

| IMD, deciles | Civil service or Local Government | | Manufacturing or construction | | Health care | | Teaching and education | |
|---|---|---|---|---|---|---|---|---|
| | Total # of participants | Participants with Long COVID, n (%) | Total # of participants | Participants with Long COVID, n (%) | Total # of participants | Participants with Long COVID, n (%) | Total # of participants | Participants with Long COVID, n (%) |
| **1 (most deprived)** | 488 | 66 (13.5) | 674 | 74 (11.0) | 950 | 140 (14.7) | 854 | 136 (15.9) |
| **2** | 820 | 110 (13.4) | 1075 | 116 (10.8) | 1343 | 195 (14.5) | 1395 | 176 (12.6) |
| **3** | 986 | 113 (11.5) | 1227 | 129 (10.5) | 1609 | 201 (12.5) | 1806 | 222 (12.3) |
| **4** | 1135 | 123 (10.8) | 1447 | 131 (9.1) | 1852 | 209 (11.3) | 2095 | 255 (12.2) |
| **5** | 1125 | 97 (8.6) | 1645 | 155 (9.4) | 2051 | 244 (11.9) | 2346 | 295 (12.6) |
| **6** | 1232 | 116 (9.4) | 1834 | 149 (8.1) | 2186 | 227 (10.4) | 2758 | 350 (12.7) |
| **7** | 1425 | 140 (9.8) | 1843 | 150 (8.1) | 2408 | 243 (10.1) | 2826 | 298 (10.5) |
| **8** | 1494 | 134 (9.0) | 2064 | 165 (8.0) | 2518 | 238 (9.5) | 3158 | 372 (11.8) |
| **9** | 1676 | 148 (8.8) | 2205 | 194 (8.8) | 2696 | 250 (9.3) | 3260 | 349 (10.7) |
| **10 (least deprived)** | 1571 | 144 (9.2) | 2033 | 190 (9.4) | 2760 | 238 (8.6) | 3517 | 338 (9.6) |
| **IMD, deciles** | **Hospitality (e.g. hotel, restaurant)** | | **Retail sector (incl. wholesale)** | | **Social care** | | | |
| | Total # of participants | Participants with Long COVID, n (%) | Total # of participants | Participants with Long COVID, n (%) | Total # of participants | Participants with Long COVID, n (%) | | |
| **1 (most deprived)** | 388 | 52 (13.4) | 776 | 105 (13.5) | 379 | 50 (13.2) | | |
| **2** | 441 | 52 (11.8) | 948 | 107 (11.3) | 484 | 57 (11.8) | | |
| **3** | 560 | 57 (10.2) | 1075 | 111 (10.3) | 498 | 58 (11.7) | | |
| **4** | 608 | 60 (9.9) | 1158 | 130 (11.2) | 583 | 72 (12.4) | | |
| **5** | 622 | 66 (10.6) | 1207 | 123 (10.2) | 522 | 57 (10.9) | | |
| **6** | 650 | 67 (10.3) | 1298 | 137 (10.6) | 600 | 71 (11.8) | | |
| **7** | 659 | 60 (9.1) | 1258 | 122 (9.7) | 579 | 51 (8.8) | | |
| **8** | 671 | 73 (10.9) | 1372 | 137 (10.0) | 564 | 77 (13.7) | | |
| **9** | 646 | 55 (8.5) | 1394 | 133 (9.5) | 539 | 57 (10.6) | | |
| **10 (least deprived)** | 644 | 52 (8.1) | 1290 | 117 (9.1) | 519 | 42 (8.1) | | |

**Table S4: Association between deprivation and experiencing Long COVID at least 4 weeks after having COVID-19, for participants residing in England.**

| IMD, deciles | N=201,799 | |
|---|---|---|
| | **Odds ratio (95% CI)** | **P value** |
| **1 (most deprived)** | 1.41 (1.29, 1.54) | <0.001 |
| **2** | 1.30 (1.20, 1.41) | <0.001 |
| **3** | 1.25 (1.15, 1.35) | <0.001 |
| **4** | 1.22 (1.13, 1.32) | <0.001 |
| **5** | 1.18 (1.10, 1.28) | <0.001 |
| **6** | 1.20 (1.11, 1.29) | <0.001 |
| **7** | 1.09 (1.01, 1.17) | 0.029 |
| **8** | 1.14 (1.06, 1.23) | <0.001 |
| **9** | 1.13 (1.05, 1.21) | 0.001 |
| **10 (least deprived)** | Reference | |

Estimates Adjusted for age, ethnicity, urban/rural, comorbid conditions, household size, country, quarter of the year, healthcare and client-facing nature of the job in the multivariable logistic regression model using the logarithm of the follow-up time as an offset term.

**Table S5: Association between deprivation and experiencing Long COVID at least 4 weeks after having COVID-19, using random-effects at country level.**

| IMD, deciles | N=201,799 | |
|---|---|---|
| | **Odds ratio (95% CI)** | **P value** |
| **1 (most deprived)** | 1.46 (1.31, 1.63) | <0.001 |
| **2** | 1.35 (1.22, 1.50) | <0.001 |
| **3** | 1.29 (1.17, 1.42) | <0.001 |
| **4** | 1.24 (1.18, 1.31) | <0.001 |
| **5** | 1.23 (1.12, 1.36) | <0.001 |
| **6** | 1.21 (1.16, 1.27) | <0.001 |
| **7** | 1.11 (1.03, 1.19) | 0.004 |
| **8** | 1.17 (1.09, 1.24) | <0.001 |
| **9** | 1.13 (1.10, 1.17) | <0.001 |
| **10 (least deprived)** | Reference | |

Estimates adjusted for age, sex, ethnicity, urban/rural, comorbid conditions, household size, healthcare and patient/client-facing nature of the job, and time (as the quarter of the year) in the multilevel logistic regression model using random-effects at country level, and the logarithm of the follow-up time as an offset term.

**Table S6: Association between deprivation and experiencing Long COVID at least 4 weeks after having COVID-19, excluding those who only had self-reported COVID-19 results.**

| IMD, deciles | N= 164,469 | |
|---|---|---|
| | OR (95% CI) | P value |
| 1 (most deprived) | 1.52 (1.37, 1.69) | <0.001 |
| 2 | 1.35 (1.22, 1.49) | <0.001 |
| 3 | 1.33 (1.21, 1.46) | <0.001 |
| 4 | 1.23 (1.12, 1.35) | <0.001 |
| 5 | 1.27 (1.16, 1.39) | <0.001 |
| 6 | 1.16 (1.06, 1.27) | 0.001 |
| 7 | 1.09 (0.99, 1.19) | 0.069 |
| 8 | 1.14 (1.05, 1.25) | 0.002 |
| 9 | 1.09 (1.00, 1.19) | 0.044 |
| 10 (least deprived) | Reference | |

Estimates adjusted for age, sex, ethnicity, urban/rural, comorbid conditions, household size, healthcare and patient/client-facing nature of the job, and time (as the quarter of the year) in the multilevel logistic regression model using random-effects at country level, and the logarithm of the follow-up time as an offset term.
